# Whole genome sequencing of ‘mutation-negative’ individuals with Cornelia de Lange Syndrome

**DOI:** 10.1101/2022.09.18.22277970

**Authors:** Morad Ansari, Mihail Halachev, David Parry, Jose L. Campos, Elston N. D’Souza, Christopher Barnett, Andrew O. M. Wilkie, Angela Barnicoat, Chirag V. Patel, Elena Sukarova-Angelovska, Katta M. Girisha, Helen V. Firth, Katrina Prescott, Louise C. Wilson, Meriel McEntagart, Rosemarie Davidson, Sally Ann Lynch, Shelagh Joss, Simon T. Holden, Wayne K. Lam, Sanjay M. Sisodiya, Andrew J. Green, Gemma Poke, Nicola Whiffin, David R. FitzPatrick, Alison Meynert

## Abstract

**Aims:** This study assesses the diagnostic utility of whole genome sequence analysis in a well-characterised research cohort of individuals referred with a clinical suspicion of Cornelia de Lange syndrome (CdLS) in whom prior genetic testing had not identified a causative variant.

**Methods:** Short read, whole genome sequencing was performed in 195 individuals from 105 families, 108 of whom were affected. 100/108 of the affected individuals had prior relevant genetic testing with no pathogenic variant being identified. The study group comprised 42 trios (affected individuals with both unaffected parents), 61 singletons (unrelated affected individuals) and two families with more than one affected individual.

**Results:** 32/105 (30.5%) unrelated probands had likely causative coding region disrupting variants. 4 loci were identified in >1 proband; *NIPBL* (10), *ANKRD11* (6), *EP300* (3), *EHMT1* (2). Single alleles were detected in the remaining genes (*EBF3, KMT2A, MED13L, NLGN3, NR2F1, PHIP, PUF60, SET, SETD5, SMC1A, TBL1XR1*). Possibly causative variants in non-coding regions of *NIPBL* were identified in four individuals. Single *de novo* variants were identified in five genes not previously reported to be associated with any developmental disorder: *ARID3A, PIK3C3, MCM7, MIS18BP1* and *WDR18*.

**Conclusions:** Clustering of *de novo* non-coding variants implicate a single uORF and a small region in intron 21 in *NIPBL* regulation. Causative variants in genes encoding chromatin-associated proteins, with no defined influence on cohesin function, appear to result in CdLS-like clinical features.

## Introduction

Cornelia de Lange syndrome (CdLS) is a severe multisystem disorder characterised by malformations of the limb and diaphragm, prenatal-onset growth failure, gastrointestinal dysfunction, neurodevelopmental problems, and characteristic facies(1). Most typical CdLS is caused by heterozygous loss-of-function mutations in the gene encoding the cohesin loader, NIPBL(2, 3). Almost all *NIPBL* mutations causing typical CdLS occur *de novo* with ∼30% being post-zygotic mosaic(4, 5). Over the last 20 years mutations in genes encoding components of the cohesin ring (*SMC1A*(6), *SMC3*(7), *RAD21*(8)) or proteins required for normal DNA-cohesin interaction (*HDAC8*(9)) have been identified in individuals with atypical forms of CdLS. More recently, individuals with a provisional diagnosis of CdLS have been reported with *de novo* mutations in genes encoding chromatin associated proteins with no direct role in cohesin function e.g. *ANKRD11, SETD5* and *KMT2A*(10).

Here we present an analysis of short-read whole genome sequencing on blood- or saliva-derived DNA to analyse a cohort of 108 affected individual from 105 families with a provisional clinical diagnosis of CdLS or a CdLS-like disorder. Almost all of these individuals had previously screened negative for mutations in known CdLS genes. The results provide further support for *NIPBL* as the dominant locus in CdLS. We have identified clustered *de novo* mutations affecting the non-coding regions of *NIPBL* and balanced and unbalanced intragenic structural variants. Causative variants disrupting the coding region were identified in 14 other genes; almost all encoding chromatin-associated proteins. We also identified single *de novo* variants in five genes without strong prior evidence of association with developmental disorders.

## Materials and Methods

### Research Participant Information

The data presented in this study are derived from DNA samples and clinical information from research participants who have consented to be involved in the CdLS study managed by the MRC Human Genetics Unit in collaboration with the CdLS Foundation of UK and Ireland (http://www.cdls.org.uk). The cohort consists of 299 affected individuals with 293 unaffected relatives. These samples are held with the consent of the families obtained using a process approved by the UK multicentre research ethics committee (MREC) for Scotland (Committee A) for whole genome sequencing (04:MRE00/19; The genetics of brain growth and development). All affected individuals have been examined by an experienced clinical geneticist. Potentially diagnostic results from the research sequencing are communicated to the referring clinicians for validation in the local genetic diagnostic laboratories.

### DNA Sequencing, alignment and variant calling

WGS sequencing of the quality checked DNA was performed at Edinburgh Genomics, University of Edinburgh. FASTQ alignment used BCBio-Nextgen (0.9.7) for bam file preparation; bwa mem (v0.7.13) aligned reads to GRCh38 reference genome employing alt, decoy and HLA sequences. Duplicated fragments were marked using samblaster (v0.1.22) and indel realignment and base recalibration was performed using GATK 3.4 to create a final gVCF file.

### Diagnostic variant filtering

We used a genome-wide approach to identify *de novo* mutations in the trio samples using both cyvcf2 (11) and VASE (https://github.com/david-a-parry/vase). All probands were also screened for plausibly causative variants in known developmental disorder genes using the G2P-VEP plugin with Ensembl VEP (12). From all the variants identified in an individual, we selected only those that are rare, predicted to be functional, and potentially relevant to developmental disorders (DD) by using the G2P plugin [doi.org/10.1038/s41467-019-10016-3] in VEP [release 90.1, doi:10.1186/s13059-016-0974-4] and the DD Gene Panel (https://www.ebi.ac.uk/gene2phenotype/downloads, accessed 11/06/2018). In short, we extracted only variants satisfying the inheritance requirements of the genes in the DD Gene Panel, with MAF in public databases < 0.0001 for monoallelic and X-linked genes and MAF < 0.005 for biallelic genes. We filtered to include only variants annotated by VEP to have one of the following consequences: stop gained, stop lost, start lost, frameshift variant, inframe insertion/deletion, missense variant, coding sequence variant, initiator codon variant, transcript ablation, transcript amplification, protein altering variant, splice donor/acceptor variant (i.e., canonical splice site) or splice region variant (i.e., either within 1-3 bases of the exon or 3-8 bases of the intron). IGV plots of each candidate variant were generated from the trio, singleton and multiplex families.

### Structural variant analysis

*De novo* structural variants were called from the bam files in each trio using a paired-end and split read method (Manta; https://github.com/Illumina/manta) and a coverage-based method (Canvas https://github.com/Illumina/canvas). Each entry in the candidate SVs was associated with an image visualising the coverage and alignment within the trio.

### Annotating variants in untranslated regions

*De novo* variants identified in the 5’untranlsated region (5’UTR) of *NIPBL* were annotated with UTRannotator (https://academic.oup.com/bioinformatics/article/37/8/1171/5905476). We also annotated all variants in ClinVar (downloaded on 30/04/2022) and gnomAD v3.1.1 within the 5’UTR as defined by the MANE Select (14) transcript (chr5:36876769-36877178 and chr5:36953618-36953696 on GRCh38). We retained all variants with an annotation indicative of creating an upstream start-codon (uAUG-gained) or disrupting a predicted upstream open reading frame (uORF; uAUG-lost, uSTOP-lost, uSTOP-gained or uFrameshift). ClinVar variants were further filtered to those classified as Pathogenic, Likely_Pathogenic, or Pathogenic/Likely_pathogenic. Finally, we searched the literature for any additional 5’UTR variants identified in individuals with CdLS. The strength of the Kozak consensus surrounding each uAUG was defined as either Weak, Moderate or Strong, as has been done previously.

### Generation of Protein Images

The R package drawProteins (13) was used to generate cartoons of the domain structure of proteins encoded by the MANE Select (14) transcript using data obtained from UniProt (15). The position of the variants predicted to affect the coding region were added using simple R commands using the R packages ggplot2 (16).

## Results

### Case selection

This study was designed to assess short-read WGS as a diagnostic tool in CdLS. Following a review of DNA quality and prior molecular genetic analysis in the 299 affected individuals participating in the MRC HGU CdLS cohort, we identified 100 affected individuals who had screened negative for mutations in the core CdLS genes (*NIPBL, SMC1A, SMC3, RAD21* and *HDAC8*) and 8 probands who had no prior screening of the CdLS genes. The available growth details and clinical synopsis relating to the affected individuals discussed below is provided in **Supplementary Table 1**. The WGS cohort consisted of 61 singletons, 42 trios and 2 quads (one affected sib pair with both unaffected parents and one affected sib pair with an affected and an unaffected parent), for a total of 195 individuals, 108 of whom being affected.

### Variants filtering

WGS reads were generated on 195 individuals and processed using the MRC Human Genetics Unit pipeline (see Methods). Since our primary aim is molecular diagnoses of affected individuals, the analyses focussed on identifying moderate or high impact rare variants that occurred; (1) using trio and quad families to identify *de novo* variants using a combination of cyvcf2 and VASE and (2) screening known developmental disorder genes in all individuals using the G2P-VEP plugin with the DDG2P dataset. We identified 60 candidate monoallelic (heterozygous or hemizygous) variants in 54 probands that survived our filtering (see methods; **Supplementary Table 2**). No plausible biallelic genotypes survived filtering. 32 variants in 32 probands were scored as pathogenic or likely pathogenic (P/LP) using ACMG criteria(18, 19). 6 variants in 4 probands were identified in the non-coding regions of *NIPBL*. 5 *de novo* variants in 5 probands were identified in genes not previously associated with developmental disorders.

### Pathogenic or Likely Pathogenic (P/LP) variants

Heterozygous loss-of-function mutations in the coding regions of *NIPBL* are, by far, the most common class of causative variant associated with CdLS (2, 3, 5, 20). We identified 10 P/LP monoallelic *NIPBL* variants in 10 different probands (**Table 1, Figure 1A, Supplementary Figures 1&2**). 8 of these could be shown to have occurred *de novo* (**Table 1**) and for two probands the parental samples were not available for testing. 9/10 represent clear loss-of-function (LOF) variants: 1 stop gain (4445), 3 frameshift (3616, 5263 and 5651), 3 essential splice site (4536, 4691 and 5320), 1 disruptive intragenic inversion (4197, **Figure 2B**) and 1 intragenic deletion removing the most 3’ coding exons (4497, **Figure 2C**). We also identified a *de novo* missense variant (p.(Ala34Val)) in proband 4281 within a region that mediates the interaction of NIPBL with MAU2 (21). *In silico* predictors (SIFT: Deleterious (0.01); PolyPhen: Probably damaging (0.98); CADD 26.2; REVEL 0.59; SpliceAI ≤ 0.2) are broadly supportive of a deleterious effect.

**Table 1:**
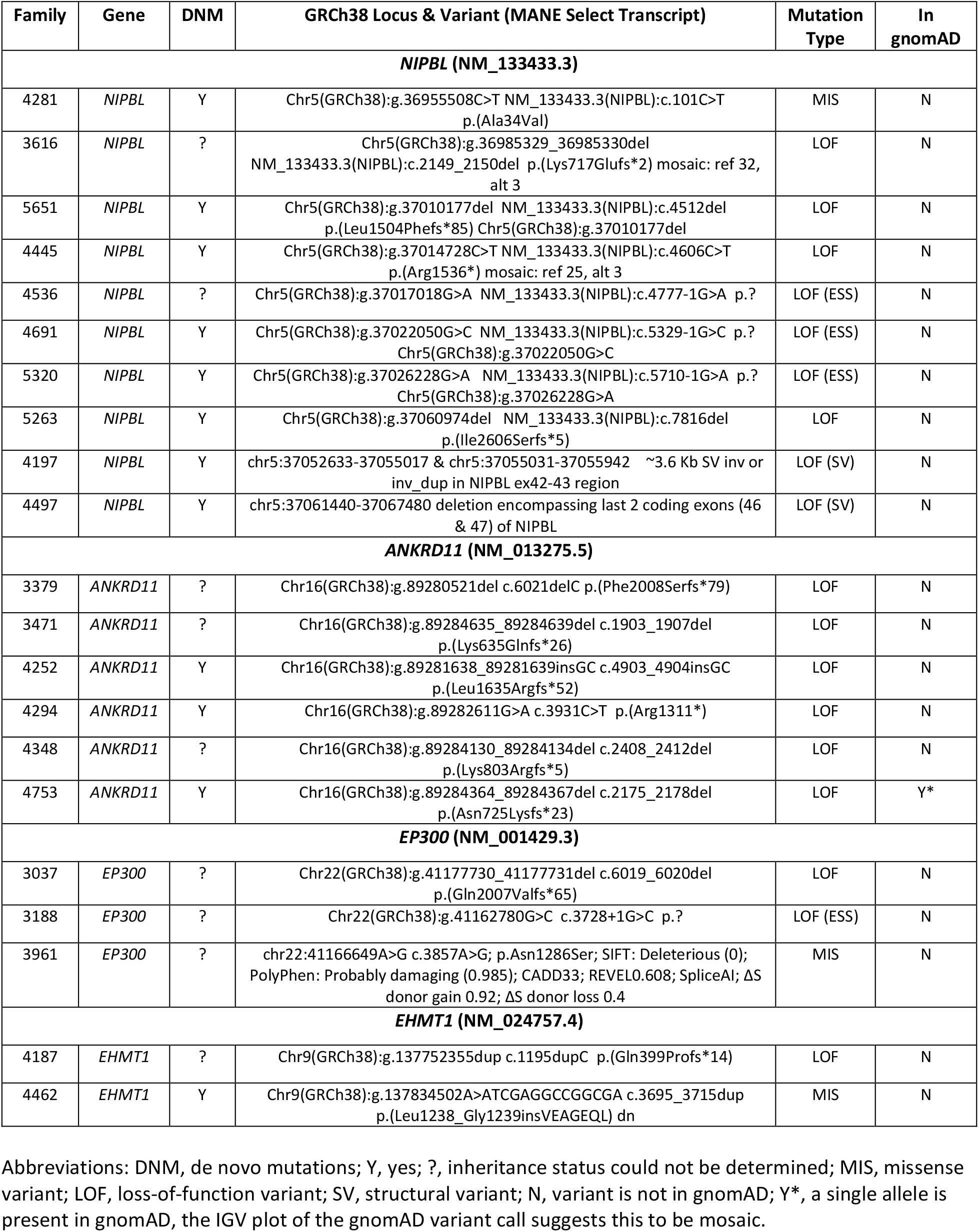
Genes with pathogenic and likely pathogenic variants in > 1 proband.

**Figure 1:**
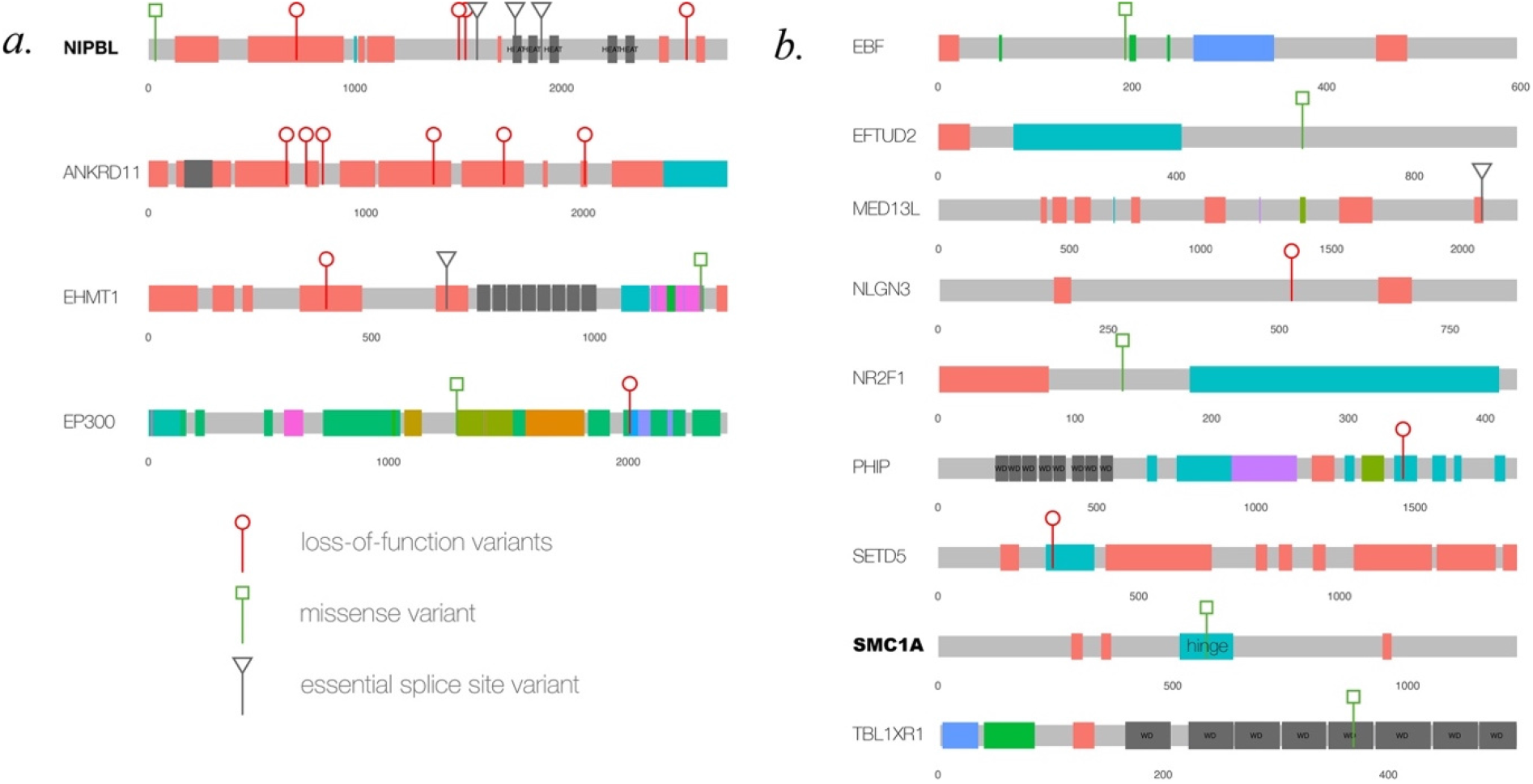
Pathogenic or likely pathogenic variant in known developmental disorder loci. This figure shows cartoons of 13 different proteins encoded by the loci in which causative heterozygous variants have been identified in this study. Each of these loci are known causes of developmental disorders. The proteins in bold script have a direct role in mediating the normal function of cohesin. A. Four protein in which variants in >1 unrelated affected individual has been identified. The position and type of the variants is indicated using the key below this panel. B. Proteins, mutation type and position of the variants that have been identified in a single proband. The domain name is indicated when a missense variant lies within the domain.

**Figure 2:**
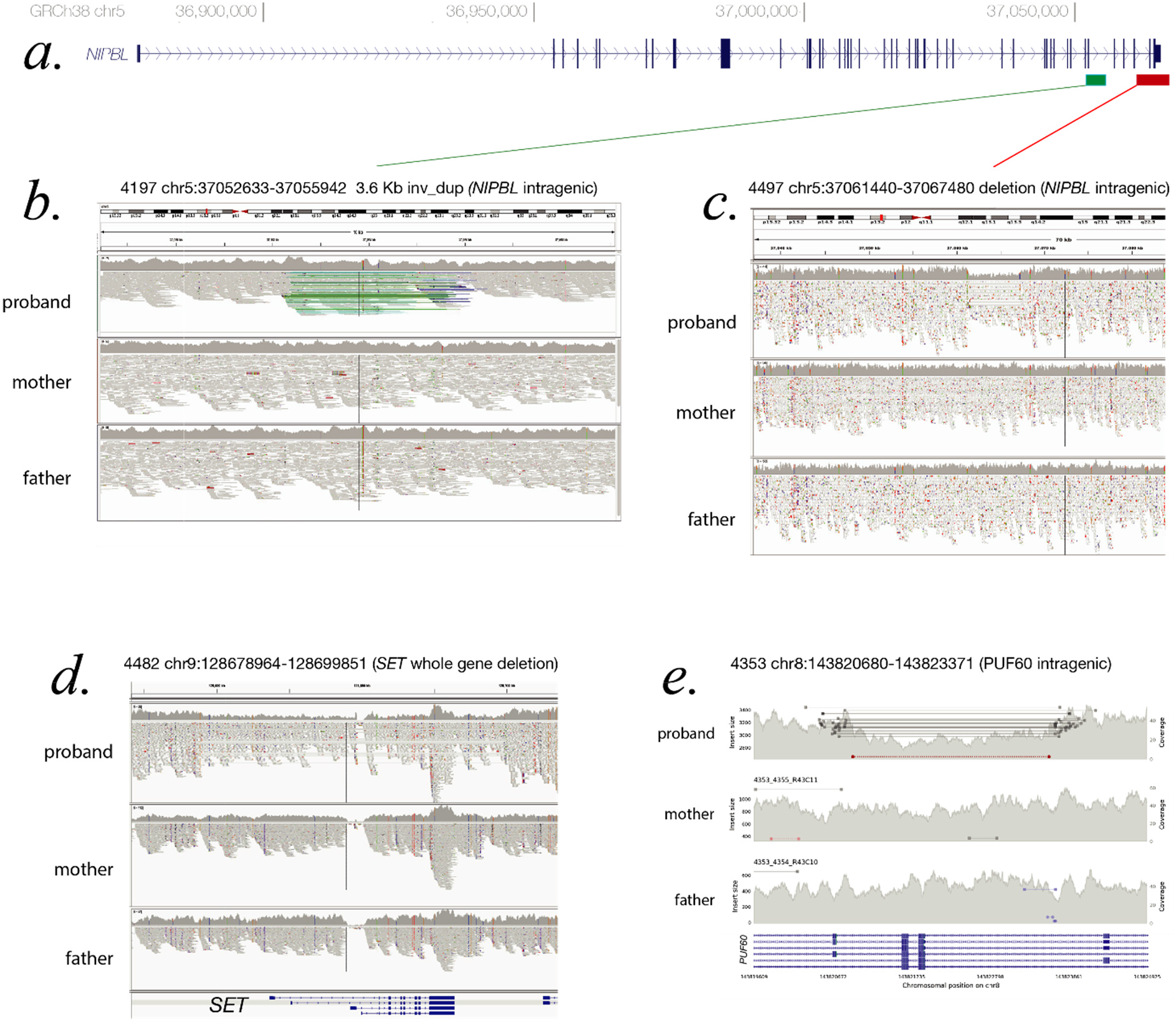
Causative structural variants. A. Cartoon of the genomic structure of *NIPBL* coloured bars indicating the position of the structural variants shown in B and C. B. IGV plot of the proband 4197 and their parents showing a region of chromosome 5. The green lines on the proband IGV plot indicate and inverted segment of chromosome with the blue lines representing a possible duplicated region (the coverage graph does not support this increased copy number). The inversion is predicted to encompass *NIPBL* exons 42 and 43 and disrupt the open reading frame. C. IGV plot of proband 4497 and their parents. A heterozygous, *de novo* deleted region is indicated by the drop in coverage in the proband and the grey lines on the IGV plot indicating paired end reads that cover the deletion breakpoints. This deletion encompasses exon 46 and 47 which encode the most C-terminal region of NIPBL. D. The IGV plot of proband 4482 and their parents indicating a de novo deletion encompassing the whole *SET* gene.

Heterozygous LOF variants in *ANKRD11* are, most commonly, associated with KBG syndrome (22, 23) but a phenotypic overlap with CdLS has been recurrently reported (5, 24–26). We identified 6 P/LP LOF variants in *ANKRD11* in 6 unrelated probands (**Table 1, Figure 1A, Supplementary Figure 3**). In three families these variants arose *de novo* (4252, 4294 and 4753) but for the remaining probands (3379, 3471 and 4348) parental samples were not available. Plausibly causative heterozygous variants in *EP300* and *EHMT1* were identified in 3 (3037, 3188 and 3961) and 2 (4187 and 4462 (de novo)) probands respectively (**Figure 1A, Table 1**). Variants in both loci have been previously reported in individuals with a clinical suspicion of CdLS (24, 27).

Single probands with P/LP variants in 11 additional genes (*EBF3, KMT2A, MED13L, NLGN3, NR2F1, PHIP, PUF60, SET, SETD5, SMC1A* and *TBL1XR1*) are documented in **Figure 1B** and **Table 2**. The *de novo* heterozygous missense variant in the hinge domain of *SMC1A* identified in proband 5661 is typical of CdLS-associated variants in this gene (6, 28–30). *KMT2A, MED13L, PHIP* and *SETD5* would not commonly be referred to as CdLS genes but the heterozygous LOF mutations identified in probands 3236, 3057, 4248 and 3036, respectively, are comparable to those previously reported in CdLS (10, 24). The remaining six probands (4021, 4482, 4383, 3046, 4353 & 3035) have variants in genes which have not been implicated in CdLS before but are known to be associated with non-syndromic (*NLGN3* (31), *SET* (32)) and/or syndromic (*EBF3* (33), *NR2F1* (34), *PUF60* (35), *TBL1XR1* (36, 37)) intellectual disability, respectively (proven *de novo* in 4482 and 4353). We could not determine whether these variants represent false positive, contributary or fully explanatory molecular diagnoses for the CdLS-like phenotype in the probands.

**Table 2:** Genes with pathogenic and likely pathogenic variants in a single proband.

| Family | Gene | DNM | Variant (GRCh38) | Mutation Type | In gnomAD |
| --- | --- | --- | --- | --- | --- |
| 4383 | *EBF3* | Y | GRCh38 10:129877825: ENST00000440978.2:c.579G>T; ENSP00000387543.2:p.Lys193Asn; SIFT: Deleterious (0); PolyPhen: Probably damaging (1); CADD28; REVEL0.437; SpliceAI≤ 0.2 | MIS | N |
| 3236 | *KMT2A* | ? | **Chr11(GRCh38):g.118484286dup NM_001197104.1(KMT2A):c.4190dup p.(Val1398Serfs\*9)** | LOF | N |
| 3057 | *MED13L* | ? | **Chr12(GRCh38):g.115966244C>A NM_015335.4(MED13L):c.6226-1G>T p.?** | LOF (ESS) | N |
| 4021 | *NLGN3* | ? | chrX:71167650G>A:HEM: NM_181303.2(NLGN3):c.1553G>A p.(Trp518*) | LOF | N |
| 3046 | *NR2F1* | ? | chr5:93585427G>A:ENST00000327111.8:c.404G>A; ENSP00000325819.3:p.Arg135His; SIFT: Deleterious (0); PolyPhen: Probably damaging (1); CADD32; REVEL0.962; SpliceAI≤ 0.2 | MIS | N |
| 4248 | *PHIP* | ? | **Chr6(GRCh38):g.78946244dup NM_017934.7(PHIP):c.4387dup p.(Arg1463Lysfs\*35) g.dupT c.dupA** | LOF | N |
| 4353 | *PUF60* | Y | chr8:143820938-143823597 deletes exons 3 and 4 PUF60 | LOF (SV) | N |
| 4482 | *SET* | Y | chr9:128678964-128699851 deletion encompassing *SET* | LOF (SV) | N |
| 3036 | *SETD5* | ? | **Chr3(GRCh38):g.9441638del NM_001080517.1(SETD5):c.856del p.(Leu286\*)** | LOF | N |
| 5661 | *SMC1A* | Y | **GRCh38 X:53405788: ENST00000322213.9:c.1714C>T; ENSP00000323421.3:p.Pro572Ser; SIFT: Deleterious (0.01); PolyPhen: Probably damaging (0.996); CADD24.8; REVEL0.86; SpliceAI≤ 0.2;** | MIS | N |
| 3053 | *TBL1XR1* | ? | chr3:177038113G>T:HET:ENST00000457928.7:c.1107C>A; ENSP00000413251.3:p.Asp369Glu; SIFT: Deleterious (0.02); PolyPhen: Probably damaging (0.985); CADD23.7; REVEL0.379; SpliceAI≤ 0.2 | MIS | N |

Abbreviations: DNM, de novo mutations; Y, yes; ?, inheritance status could not be determined; MIS, missense variant; LOF, loss-of-function variant; SV, structural variant; N, variant is not in gnomAD. The rows in bolded text are loci that have previously been reported to be implicated in the pathogenesis of CdLS.

### Clustering of non-coding variants in *NIPBL*

We identified two probands with *de novo* variants in the first exon of *NIPBL* (4079 & 4709; **Table 3, Figure 3A,B**) which encodes part of the 5’UTR. The 5’UTR of *NIPBL* contains five predicted uORFs, three within exon 1 (**Figure 3A**). The *de novo* variant in proband 4079 (c.-467C>T) creates a novel upstream start codon (uAUG) into a strong Kozak consensus context, creating a new uORF that is 156bps in length (**Figure 3A**). This variant was previously identified *de novo* in an individual with CdLS. Interestingly, two further variants reported in the literature are predicted to also create uAUGs: (1) the c.-457_-456delinsAT variant identified *de novo* in a 15-year-old male with classic CdLS (moderate Kozak; 270bp long uORF created), and (2) the c.-94C>T variant which creates a uAUG with a weak match to the Kozak consensus in a patient with a mild phenotype (**Figure 3A**). The *de novo* variant in proband 4709 (c.-315del) has not been observed previously. This variant deletes a single base of the 5’UTR directly following the uAUG of an existing uORF with a moderate Kozak match. The variant shifts the reading frame of the uORF extending it from 15bps to 189bps in length (**Figure 3A, Supplementary Table 3**). A different 5’UTR variant reported previously (c.-321_-320delinsA) has the same predicted impact. We searched the gnomAD v3.1.1 dataset for 5’UTR variants with similar predicted effects (**Supplementary Table 4**). Whilst two variants, each identified in a single gnomAD individual, create uAUGs, both have a weak match to the Kozak consensus. Six variants are predicted to shift the frame of an existing uORF, but the impacted uORFs also have a weak Kozak consensus so are unlikely to be strongly translated. The clustering and predicted consequence of 5’UTR variants in CdLS patients suggests an important role for uORF regulation in NIPBL translation.

**Figure 3:**
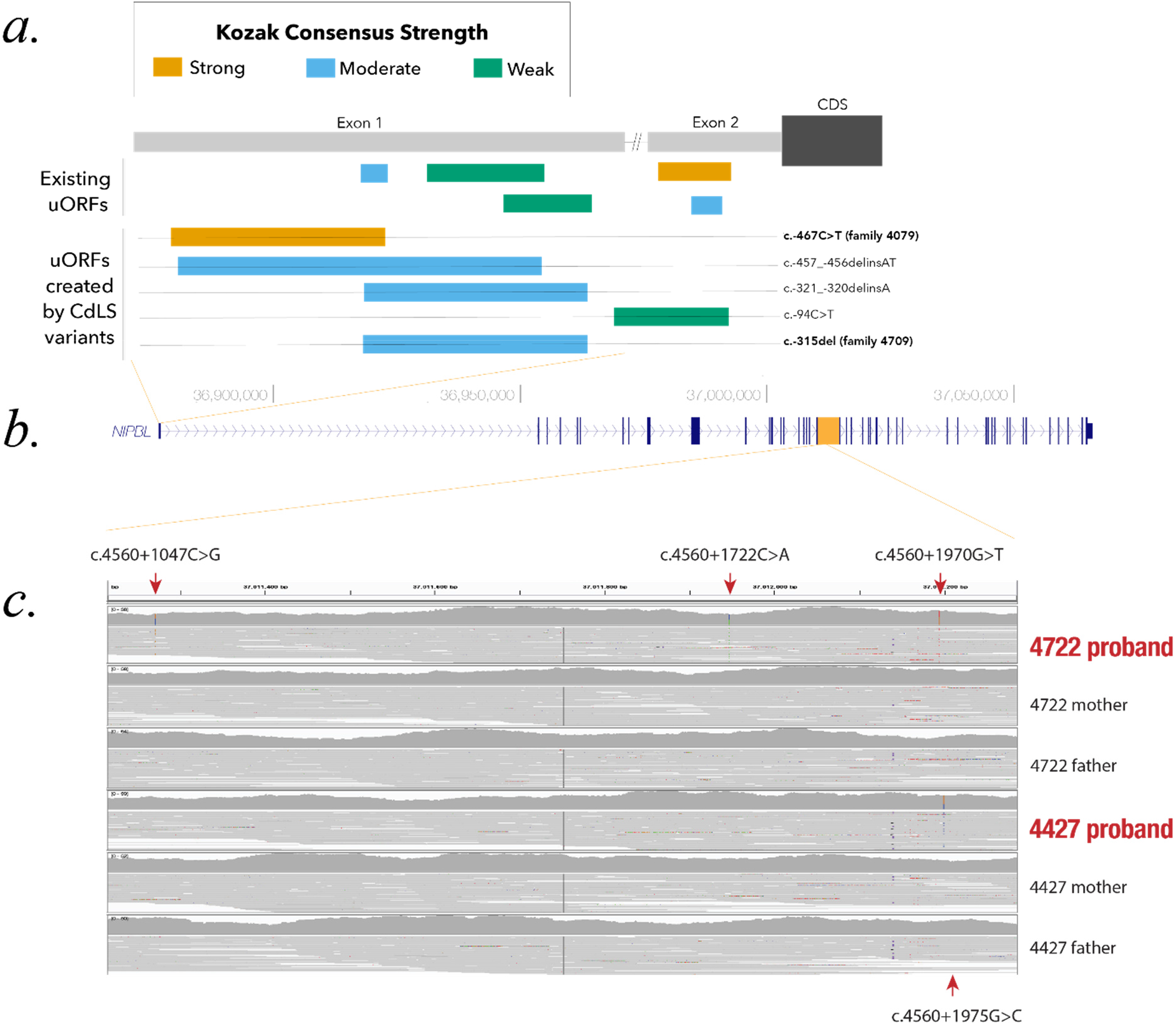
De novo variants affecting uORF structure and clustering in intron 21 of *NIPBL*. A. Cartoon of the position of the predicted uORFs in the 5’UTR encoded by exon1 and exon 2 of *NIPBL*, indicating the strength of the Kozak translational start sequence shown in yellow, blue and green for strong, moderate and weak, respectively. The positions of the de novo variants in probands 4079 and 4709 and their predicted effects are also shown. B. Cartoon of the NIPBL genomic structure derived from the UCSC Genome Browser indicating the position of the non-coding variants detailed in A and C. C. IGV snapshot of the ∼1kb interval containing the de novo, deep intronic variants identified in Intron 21. Three de novo variants (arrowed above the IGV plots) were identified in proband 4722 and a single variant (arrowed below the IGV plot) in proband 4427.

**Table 3:** *De novo* non-coding variants in *NIPBL*.

| Family | Gene | DNM | GRCh38 coordinates | cDNA<br>(NM_133433.4) | CADD | SpliceAI | Mutation<br>Type | In<br>gnomAD |
| --- | --- | --- | --- | --- | --- | --- | --- | --- |
| 4079 | *NIPBL* | Y | 5:36876791C>T | c.-467C>T | 20.1 | ≤ 0.2 | NC (uORF) | N |
| 4709 | *NIPBL* | Y | 5:36876943del | c.-315del | 19.7 | ≤ 0.2 | NC (uORF) | N |
| 4427 | *NIPBL* | Y | 5:37012200G>C | c.4560+1975G>C | 1.3 | ≤ 0.2 | NC (int 21) | N |
| 4722 | *NIPBL* | Y | 5:37011272C>G | c.4560+1047C>G | 0.6 | ≤ 0.2 | NC (int 21) | N |
|  |  | Y | 5:37011947C>A | c.4560+1722C>A | 4.3 | ≤ 0.2 | NC (int 21) | N |
|  |  | Y | 5:37012195G>T | c.4560+1970G>T | 1.5 | ≤ 0.2 | NC (int 21) | N |

Abbreviations: DNM, de novo mutations; Y, yes; NC, non-coding variant; uORF, upstream open reading frame in 5'UTR; int 21, intron 21 of *NIPBL* gene; N, variant is not in gnomAD.

In proband 4722 we identified three different *de novo* variants within a 1kb region of Intron 21 (**Table 3, Figure 3C**). In proband 4427 we identified a single *de novo* variant (c.4560+1975G>C, **Table 3, Figure 3C**) that is only 5 base pairs away from the most 3’ 4722 variant (c.4560+1970G>T) within a SINE repeat element. None of these deep intronic variants are in gnomAD, none show evidence of a deleterious effect on splicing and each has a low CADD score (**Table 3**). We are currently unable to perform any functional analysis of this segment of intron 21 and thus cannot predict a consequence for these variants.

### *De novo* variants in genes not previously implicated in developmental disorders

Following the IGV inspection of candidate de novo calls, 5 variants in 5 “novel” genes (i.e. not present in the DDG2P dataset) were identified in 5 different probands (**Table 4, Figure 4A**) including individual 4353 who also has a *de novo* intragenic deletion in *PUF60* (**Table 2, Figure 2**) making it difficult to attribute any contribution of *MIS18BP1* to the phenotype. In proband 4954, in silico predictions show only weak evidence of deleteriousness for the *WDR18* missense variant. Neither of these variants will be considered further. Of the remaining genes (*PIK3C3, MCM7, ARID3A*) only *MCM7* (proband 4485) has any direct link to cohesin function. *MCM7* encodes a subunit of the replicative helicase MCM2-7 which is required for the loading of cohesin onto DNA during S-phase. *ARID3A* encodes a widely expressed transcription factor with roles in haematopoiesis, placental development, and mesoderm formation. *PIK3C3* encodes a component of the complex that catalyses phosphatidylinositol 3-phosphate formation. Mechanistically, this would not represent an obvious candidate gene for CdLS.

**Table 4:** *De novo* variants in genes not known to cause developmental disorders.

| Family | Gene | DNM | Variant(s) of note | Mutation Type | In gnomAD |
| --- | --- | --- | --- | --- | --- |
| 3060 | *PIK3C3* | Y | 18:41957625C>T; ENST00000262039.9:c.124C>T; ENSP00000262039.3:p.Pro42Ser; SIFT: Deleterious (0); PolyPhen: Probably damaging (0.971); CADD 25.4; REVEL 0.66; SpliceAI ≤ 0.2 | MIS | N |
| 4353* | *MIS18BP1* | Y | 14:45226740-45226750<br>GRCh38 (NM_018353.4):c.1833_1840+3delinsAACC, p.(Lys612Thrfs*14) | LOF | N |
| 4485 | *MCM7\*\** | Y | 7: 100098712G>A ENST00000303887.10<br>c.586C>T p.Gln196Ter | LOF | N |
| 4847 | *ARID3A* | Y | 19:964425G>A: ENST00000263620.8:c.944G>A; ENSP00000263620.2:p.Arg315Gln; SIFT: Deleterious (0.02); PolyPhen: Probably damaging (0.993); CADD 32; REVEL 0.72; SpliceAI ≤ 0.2 | MIS | N |
| 4954 | *WDR18* | Y | 19:991291: ENST00000585809.6:c.871G>A; ENSP00000476117.3:p.Glu291Lys; SIFT: Tolerated (0.12); PolyPhen: Possibly damaging (0.498); CADD 25.6; REVEL 0.268; SpliceAI ≤ 0.2 | MIS | N |

Abbreviations: DNM, de novo mutations; Y, yes; MIS, missense variant; LOF, loss-of-function variant; SV, structural variant; N, variant is not in gnomAD. \*This proband also has an intragenic PUF60 deletion (see Table 2); \*\* This variant may be mosaic in the proband with ref:alt ratio 30:10.

**Figure 4.**
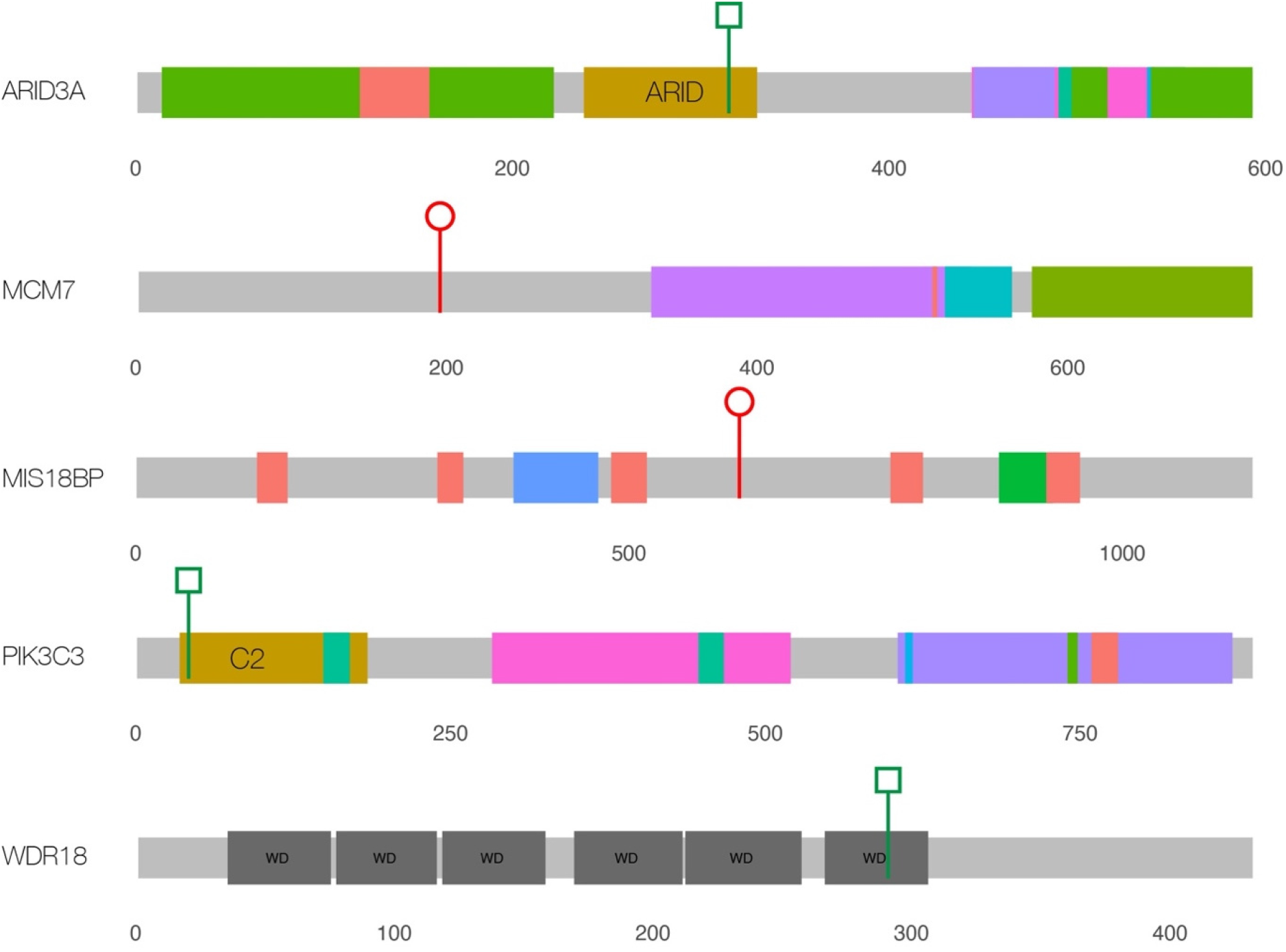
De novo protein coding variants in genes not known to be associated with developmental disorders. This figure shows cartoons of 5 different proteins encoded by the loci in which de novo variants have been identified in this study. None of these loci are known causes of developmental disorders.

## Discussion

Diagnostic genomic analysis of individuals with severe developmental disorders can confidently identify genes with an important and non-redundant developmental role. It is reasonable to hypothesise that the identification of these products will indicate specific critical functions they mediate during embryogenesis and improve our understanding of the developmental pathology. CdLS is very commonly described as a cohesinopathy(40) on the grounds of the phenotypic overlap of individuals with mutations in genes encoding the components of the cohesin ring and factors regulating its interaction with DNA. However, the very large number of different functions of cohesin somewhat limits our understanding of the specific disease mechanisms. It is not unreasonable to assume that identifying other disease loci with significant phenotypic overlap with CdLS may implicate perturbation of a subset of cohesin roles in the disease mechanism.

In every published CdLS cohort analysis, *NIPBL* is by far the most frequently mutated gene (3, 4, 24, 41–44). We have previously reported a screen of a cohort of 168 individuals enriched with atypical CdLS(5). 63/168 (37.5%) had coding region mutations in the known CdLS genes (*NIPBL, SMC1A, SMC3, RAD21, HDAC8*) with 75% of the causal variants affecting the coding region of *NIPBL*. Given the almost universal association of severe typical CdLS with *NIPBL*, we estimated that a further ∼20% of the unexplained cases are likely to be due to cryptic mutations or mosaicism at this locus. The current study was not designed to detect mosaicism as it was based predominantly on the analysis of blood-derived DNA. However, one of the main advantages of diagnostic WGS is the identification of plausibly pathogenic variants in the non-coding regions of the transcription unit that would be missed on most WES analyses. In this regard, the two *de novo* variants identified in the 5’UTR are particularly significant. Both have a plausible deleterious effect on translation (45), with predicted impacts similar to previously identified variants in the same region, suggesting that they are likely the causative variants in these individuals. Notably, these variants are >300 bp upstream of the start of the *NIPBL* coding sequence and would not be captured using WES. Our analysis confirms an important role for uORF regulation of *NIPBL* in CdLS suggesting that routine screening of the 5’UTR is warranted in CdLS patients. The clustered de novo deep intronic variants that we identified in intron 21 in two affected individuals are equally interesting but completely inexplicable from a mechanistic perspective. These have no predicted effect on splicing and alter bases that show no evolutionary conservation and for the most clustered variant, lie within a SINE repeat (AluJb chr5:37012140-37012330, GRCh38). This region does show TOBIAS-corrected evidence of accessibility in inner cell mass cells derived from human embryos (46, 47) but we have no other direct evidence of cis-regulatory function. We do feel that these variants should be considered “of interest” but cannot yet be considered diagnostic.

In the same cohort analysis mentioned above(5) we also identified 3 individuals with heterozygous LOF mutations in *ANKRD11* who were, from a clinical perspective, no less typical than those with mutations in *HDAC8, RAD21* and *SMC3*. Since then, many other loci have been reported as rarely causal in CdLS; *KMT2A*(24, 48), *SETD5*(24, 49), *EP300*(27), *MED13L*(24), *PHIP*(24), *AFF4*(50), *TAF6*(51), *MAU2*(52), *EHMT1*(24) and *BRD4*(53). In our current study we provide further support for the association of CdLS-like features with *ANKRD11, EP300, EHMT1, SETD5, MED13L* and *PHIP* (**Figure 1A**,**B**). Additionally, we have identified P/LP variants in *EBF, EFTUD2, NLGN3, NR2F1, TBL1XR1* (**Figure 1B**) and *SET* (**Figure 2**) in known developmental disorder loci. Most of these genes encode chromatin-associated proteins but, except for MAU2 and BRD4, they provide no evidence of direct interaction with the cohesin system. Of the genes with *de novo* variants without known disease association, only *MCM7* encodes a protein with a direct link to cohesin. We have not yet found a satisfactory unifying explanation for the CdLS-like phenotypes that are associated with this set of genes. The general term transcriptomopathy (51) is useful conceptually but, like cohesinopathy, is too broad for detailed mechanistic use.

Further analysis of the mutation-negative cases with CdLS should, ideally, exclude post-zygotic mosaic variants in *NIPBL* using analysis of DNA from a tissue such as uncultured skin. This would allow us to identify any false association in the existing data. There is a need for further experimental work focussed on identifying a functional link between NIPBL and the proteins encoded by the genes that have been recurrently identified in individuals with CdLS, most notably, ANKRD11.

## Data Availability

All data produced in the present study are available upon reasonable request to the authors

## Acknowledgements

DRF is funded by the program within the MRC University Unit award to the University of Edinburgh for the MRC Human Genetics Unit. The whole genome sequencing in this project was funded by the Simons Initiative for the Developing Brain (R83729). NW is supported by a Sir Henry Dale Fellowship jointly funded by the Wellcome Trust and the Royal Society (Grant Number 220134/Z/20/Z). NW and END are supported by grant funding from The Rosetrees Trust (Grant Number H5R01320). AOMW was supported by the National Institute for Health Research (NIHR) Oxford Biomedical Research Centre (BRC). The views expressed are those of the author(s) and not necessarily those of the NHS, the NIHR, or the Department of Health.

