## Supplementary Table 1 for "Whole genome sequencing of ‘mutation-negative’ individuals with Cornelia de Lange Syndrome"

**Supplementary Table 1 - CdLS categories**

| Proband ID | Family ID | CdLS Classification | Height_SD | Weight_SD | OFC_SD |
|---|---|---|---|---|---|
| 3027 | 3027 | Mild | -2.27 | 0.01 | -0.47 |
| 3028 | 3028 | Mild | -1.15 | -0.94 | -1.21 |
| 3028 | 3028 | Mild | -0.35 | | -1.49 |
| 3036 | 3036 | Possible | | | |
| 3037 | 3037 | Atypical | | | |
| 3040 | 3040 | Possible | | | |
| 3041 | 3041 | Typical | | | |
| 3046 | 3046 | Possible | | | |
| 3053 | 3053 | Typical | -3.01 | -3.03 | -2.26 |
| 3057 | 3057 | Possible | -1.98 | 0.35 | 0.27 |
| 3060 | 3060 | Possible | | | |
| 3177 | 3177 | Typical | | | |
| 3188 | 3188 | Possible | | | |
| 3236 | 3236 | Typical | | -2.32 | |
| 3379 | 3379 | Possible | -2.29 | -2.31 | |
| 3461 | 3461 | Possible | 0.14 | 1.64 | -3.68 |
| 3471 | 3471 | Possible | | | -0.26 |
| 3616 | 3616 | Atypical | -0.84 | -0.77 | -3.93 |
| 3778 | 3778 | Uncertain | | | |
| 3961 | 3961 | Atypical | -2.49 | -2.55 | -5.05 |
| 4021 | 4021 | Atypical | | | |
| 4075 | 4075 | Typical | | -4.93 | -5.29 |
| 4079 | 4079 | Possible | -4.04 | | -4.76 |
| 4187 | 4187 | Possible | | | |
| 4197 | 4197 | Typical | -4.36 | -5.18 | -7.24 |
| 4252 | 4252 | Possible | | -3.06 | -0.18 |
| 4252 | 4252 | Uncertain | -3.43 | -3.03 | |
| 4281 | 4281 | Possible | | | |
| 4294 | 4294 | Possible | | | -0.8 |
| 4306 | 4306 | Typical | | | |
| 4348 | 4348 | Typical | | | |
| 4353 | 4353 | Mild | -1.11 | -0.03 | -2.64 |
| 4383 | 4383 | Possible | -3.02 | -2.73 | -0.61 |
| 4427 | 4427 | Mild | -3.4 | -2.02 | -2.31 |
| 4441 | 4441 | Possible | | | |
| 4445 | 4445 | Typical | | | -4.21 |
| 4462 | 4462 | Typical | -4.45 | -9.91 | -5.42 |
| 4485 | 4485 | Typical | -1.18 | -2.58 | -2.66 |
| 4497 | 4497 | Typical | -3.7 | -2.76 | -3.11 |
| 4536 | 4536 | Uncertain | | | |
| 4665 | 4665 | Mild | -3.63 | -1.67 | -2 |
| 4691 | 4691 | Typical | | | -4.87 |

| | | | | | |
|---|---|---|---|---|---|
| 4709 | 4709 | Atypical | | | |
| 4722 | 4722 | Atypical | -1.5 | -2.74 | -3.78 |
| 4753 | 4753 | Atypical | 0.6 | -0.53 | 0.22 |
| 5263 | 5263 | Possible | -4.83 | -8.07 | -6.93 |
| 5320 | 5320 | Typical | | -5.29 | |
| 5651 | 5651 | Atypical | | -3.63 | -4.87 |
| 5661 | 5661 | Typical | | | |
