## Supplementary Table 2 for "Whole genome sequencing of ‘mutation-negative’ individuals with Cornelia de Lange Syndrome"

**Supplementary Table 2 - Candidate monoallelic (heterozygous or hemizygous) variants that survived filtering**

| Family | Gene | DDG2P | De novo | Variant(s) of note | Amino_Acid_number | Mutation_Type | In gnomAD | DRF | Pathogenicity prediction |
|---|---|---|---|---|---|---|---|---|---|
| 3053 | TBL1XR1 | Y | ? | chr3:177038113:177038113:G:T:HET:ENST00000457928.7:c.1107C>A; ENSP00000413251.3:p.Asp369Glu; SIFT: Deleterious (0.02); PolyPhen: Probably damaging (0.985); CADD23.7; REVEL0.379; SpliceAI≤ 0.2 | 369 | MIS | N | DDD-WOS / Patient 260528 in cluster of LP/P missense see PMID: 26740553 | Likely Pathogenic |
| 4079 | NIPBL | Y | Y | 5:36876791C>T:ENST00000282516.13:c.-467C>T; CADD20.1; SpliceAI≤ 0.2 | | NC | N | De novo | Possible |
| 4197 | NIPBL | Y | Y | chr5:37052633-37055017 chr5:37055031-37055942 ~3.6 Kb SV inv or inv_dup in NIPBL ex42-43 region | | DEL | N | De novo | Likely Pathogenic |
| 3043 | KCNQ2 | Y | ? | chr20:63414092:63414092:C:T:HET:ENST00000359125.7:c.1627G>A; ENSP00000352035.2:p.Val543Met; SIFT: Deleterious (0.02); PolyPhen: Probably damaging (0.936); CADD25; REVEL0.774; SpliceAI≤ 0.2 | | | Y | No seizures seems unlikely | Not LP/P |
| 3043 | POLD1 | Y | ? | chr19:50402637:50402637:A:G:ENST00000440232.7:c.866A>G; ENSP00000406046.1:p.Asp289Gly; SIFT: Deleterious (0.01); PolyPhen: Probably damaging (0.925); CADD26.4; REVEL0.413; SpliceAI≤ 0.2 | | | Y | Unlikely | Not LP/P |
| 4281 | NIPBL | Y | Y | Chr5(GRCh38):g.36955508C>T NM_133433.3(NIPBL):c.101C>T p.(Ala34Val) | 34 | MIS | N | De novo | Pathogenic |
| 3052 | MYT1 | Y | ? | Chr20(GRCh38):g.64212121C>G ENST00000328439.6:c.1500C>G; ENSP00000327465.1:p.Asn500Lys; SIFT: Deleterious (0); PolyPhen: Probably damaging (0.992); CADD26.1; REVEL0.245; SpliceAI≤ 0.2 | | | N | REVEL and CADD not strong, no clinical fit | Not LP/P |
| 4353 | PUF60 | Y | Y | chr8:143820938-143823597 deletes exons 3 and 4 PUF60 | | DEL | N | De novo | Likely Pathogenic |
| 4427 | NIPBL | Y | Y | GRCh38 5:37012200-37012200: ENST00000282516.13:c.4560+1975G>C; CADD1.325; SpliceAI≤ 0.2 | | NC | N | De novo | Possible |
| 4445 | NIPBL | Y | Y | Chr5(GRCh38):g.37014728C>T NM_133433.3(NIPBL):c.4606C>T p.(Arg1536*) mosaic: ref 25, alt 3 | 1536 | LOF | N | De novo | Pathogenic |
| 3062 | DDX3X | Y | ? | chrX:41341583:41341583:T:G:HEM:ENST00000644876.2:c.251T>G; ENSP00000494040.1:p.Phe84Cys; SIFT: Tolerated (0.15); PolyPhen: Benign (0); CADD22.9; REVEL0.16; SpliceAI≤ 0.2 | | | Y | No support | Not LP/P |
| 4462 | EHMT1 | Y | Y | NM_024757.4(EHMT1):c.3695_3715dup p.(Leu1238_Gly1239insVEAGEQL) dn | 1238 | MIS | N | De novo | Pathogenic |
| 4497 | NIPBL | Y | Y | last 2 or 3 exons (exons 45-47, del of approx. 37061000-37067000) and downstream 5:37072385>G>C | | DEL | N | De novo | Pathogenic |
| 4691 | NIPBL | Y | Y | Chr5(GRCh38):g.37022050G>C NM_133433.3(NIPBL):c.5329-1G>C p.? Chr5(GRCh38):g.37022050G>C | 1776 | ESS | N | De novo | Pathogenic |
| 4709 | NIPBL | Y | Y | 5:36876936-36876937del: ENST00000282516.13:c.-321del 5_prime_UTR_variant SpliceAI≤ 0.2 | | NC | N | De novo | Possible |
| 3461 | NUP210 | N | Y | GRCh38 3:13360354-13360354: ENST00000254508.7:c.2070C>G; ENSP00000254508.5:p.Ile690Met; SIFT: Deleterious (0.01); PolyPhen: Possibly damaging (0.533); CADD17.58; REVEL0.047; SpliceAI≤ 0.2 | | | N | De novo, no support | Not LP/P |
| 4722 | NIPBL | Y | Y | Chr5(GRCh38):g.37011272C>G NM_133433.4(NIPBL):c.4560+1047C>G AND Chr5(GRCh38):g.37011947C>A NM_133433.4(NIPBL):c.4560+1722C>A AND Chr5(GRCh38):g.37012195G>T NM_133433.4(NIPBL):c.4560+1970G>T Intron 21 | | NC | N | De novo | Possible |
| 3471 | ARID1B | Y | ? | chr6:156778617:156778617:T:G:ENST00000636930.2:c.937T>G; ENSP00000490491.2:p.Phe313Val; SIFT: Deleterious low confidence (0); PolyPhen: Probably damaging (0.997); CADD23.2; REVEL0.051; SpliceAI≤ 0.2 | | | N | REVEL and CADD not strong | Not LP/P |
| 5263 | NIPBL | Y | Y | Chr5(GRCh38):g.37060974del NM_133433.3(NIPBL):c.7816del p.(Ile2606Serfs*5) | 2606 | LOF | N | De novo | Pathogenic |

| | | | | | | | | | |
|---|---|---|---|---|---|---|---|---|---|
| 3617 | STAG1 | Y | ? | chr3:136472477:136472477:T:C:ENST00000383202.7:c.1141A>G; ENSP00000372689.2:p.Met381Val; SIFT: Deleterious (0); PolyPhen: Possibly damaging (0.601); CADD25.6; REVEL 0.721; SpliceAI≤ 0.2 | 381 | MIS | N | Inheritance unknown, close to a cluster of pathogenic missense variants | Uncertain |
| 3778 | EMC1 | Y | ? | chr1:19243668:19243668:G:A:ENST00000477853.6:c.326C>T; ENSP00000420608.1:p.Ser109Phe; SIFT: Deleterious (0); PolyPhen: Probably damaging (0.999); CADD31; REVEL0.316; SpliceAI≤ 0.2 | 109 | MIS | N | Inheritance unknown, monoallelic varants reported but with cerebellar atrophy as major component, REVEL not strong but CADD high | Uncertain |
| 5320 | NIPBL | Y | Y | Chr5(GRCh38):g.37026228G>A NM_133433.3(NIPBL):c.5710-1G>A p.? Chr5(GRCh38):g.37026228G>A | 1903 | ESS | N | De novo | Pathogenic |
| 4021 | NEDD4L | Y | ? | chr18:58323243:58323243:G:A:ENST00000400345.8:c.422G>A; ENSP00000383199.2:p.Arg141Gln; SIFT: Deleterious (0.01); PolyPhen: Possibly damaging (0.729); CADD29.3; REVEL0.252; SpliceAI≤ 0.2 | | | Y | Inheritance unknown, associated with PVNH | Not LP/P |
| 5651 | NIPBL | Y | Y | Chr5(GRCh38):g.37010177del NM_133433.3(NIPBL):c.4512del p.(Leu1504Phefs*85) Chr5(GRCh38):g.37010177del | 1504 | LOF | N | De novo | Pathogenic |
| 4044 | SCN11A | Y | ? | chr3:38847388:38847388:A:T:ENST00000302328.9:c.4682T>A; ENSP00000307599.3:p.Leu1561Gln; SIFT: Deleterious (0.03); PolyPhen: Probably damaging (0.995); CADD27.3; REVEL0.879; SpliceAI≤ 0.2 | | | N | Inheritance unknown, this gene associated with episodic pain | Not LP/P |
| 5661 | SMC1A | Y | Y | GRCh38 X:53405788-53405788: ENST00000322213.9:c.1714C>T; ENSP00000323421.3:p.Pro572Ser; SIFT: Deleterious (0.01); PolyPhen: Probably damaging (0.996); CADD24.8; REVEL0.86; SpliceAI≤ 0.2; | 572 | MIS | N | De novo | Pathogenic |
| 4847 | ARID3A | N | Y | GRCh38 19:964425-964425: ENST00000263620.8:c.944G>A; ENSP00000263620.2:p.Arg315Gln; SIFT: Deleterious (0.02); PolyPhen: Probably damaging (0.993); CADD32; REVEL0.72; SpliceAI≤ 0.2 | 315 | MIS | N | De novo, high pLi, no other reports | Possible novel locus |
| 4954 | WDR18 | N | Y | GRCh38 19:991291-991291: ENST00000585809.6:c.871G>A; ENSP00000476117.3:p.Glu291Lys; SIFT: Tolerated (0.12); PolyPhen: Possibly damaging (0.498); CADD25.6; REVEL0.268; SpliceAI≤ 0.2 | 291 | MIS | N | De novo, high pLi, no other reports | Possible novel locus |
| 4353 | MIS18BP1 | Y | Y | (NM_018353.4):c.1833_1840+3delinsAACC, p.(Lys612Thrfs*14) | | | N | De novo, no disease association with this gene | Possible novel locus |
| 4485 | MCM7 | N | Y | 7:100098712-100098712 GRCh38 chr7 g.100098712G>A ENST00000303887.10 c.586C>T p.Gln196Ter mosaic Looks real: ref G 30, alt A 10 | 196 | LOF | N | De novo, no disease association with this gene, pLi 0 | Possible novel locus |
| 3060 | PIK3C3 | N | Y | GRCh38 18:41957625-41957625C>T; ENST00000262039.9:c.124C>T; ENSP00000262039.3:p.Pro42Ser; SIFT: Deleterious (0); PolyPhen: Probably damaging (0.971); CADD25.4; REVEL0.66; SpliceAI≤ 0.2 | 42 | MIS | N | De novo, no strong support PMID: 24038936, PMID: 27607605 | Possible novel locus |
| 4383 | EBF3 | Y | Y | GRCh38 10:129877825-129877825: ENST00000440978.2:c.579G>T; ENSP00000387543.2:p.Lys193Asn; SIFT: Deleterious (0); PolyPhen: Probably damaging (1); CADD28; REVEL0.437; SpliceAI≤ 0.2 | 193 | MIS | N | De novo, recurrent mutation | Pathogenic |
| 4306 | EP300 | Y | ? | chr22:41126006:41126006:A:G:HET:ENST00000263253.9:c.872A>G; ENSP00000263253.7:p.Lys291Arg; SIFT: Tolerated (0.42); PolyPhen: Probably damaging (0.948); CADD22.7; REVEL0.318; SpliceAI≤ 0.2 | | | Y | Inheritance unknown, missense predictions not supportive | Not LP/P |
| 3057 | MED13L | Y | ? | Chr12(GRCh38):g.115966244C>A NM_015335.4(MED13L):c.6226-1G>T p.? | 2075 | ESS | N | DECIPHER 291493 | Pathogenic |
| 3616 | NIPBL | Y | ? | Chr5(GRCh38):g.36985329_36985330del NM_133433.3(NIPBL):c.2149_2150del p.(Lys717Glufs*2) mosaic: ref 32, alt 3 | 717 | LOF | N | Inheritance unknown | Pathogenic |
| 4187 | EHMT1 | Y | ? | Chr9(GRCh38):g.137752355dup NM_024757.4(EHMT1):c.1195dupC p.(Gln399Profs*14) | 399 | LOF | N | Inheritance unknown | Pathogenic |
| 4536 | NIPBL | Y | ? | Chr5(GRCh38):g.37017018G>A NM_133433.3(NIPBL):c.4777-1G>A p.? | 1592 | ESS | N | Inheritance unknown | Pathogenic |

| | | | | | | | | | |
|---|---|---|---|---|---|---|---|---|---|
| 4414 | ASXL1 | Y | ? | Chr20(GRCh38):g.32434844C>T ENST00000375687.10:c.2132C>T; ENSP00000364839.4:p.Thr711Ile; SIFT: Tolerated low confidence (0.12); PolyPhen: Benign (0.029); CADD8.717; REVEL0.073; SpliceAI≤ 0.2 | | | Y | Inheritance unknown | Not LP/P |
| 3961 | EP300 | Y | ? | chr22:41166649:41166649:A:G:ENST00000263253.9:c.3857A>G; ENSP00000263253.7:p.Asn1286Ser; SIFT: Deleterious (0); PolyPhen: Probably damaging (0.985); CADD33; REVEL0.608; SpliceAI; **ΔS donor gain 0.92**; ΔS donor loss 0.4 | 1286 | MIS | N | Inheritance unknown, likely splice variant | Likely Pathogenic |
| 4441 | EFTUD2 | Y | ? | chr17:44859931:44859931:T:G:HET:ENST00000426333.7:c.1834A>C; ENSP00000392094.1:p.Lys612Gln; SIFT: Deleterious (0.01); PolyPhen: Probably damaging (0.973); CADD28.7; REVEL0.78; SpliceAI≤ 0.2 | 612 | MIS | N | Inheritance unknown, microcephaly noted | Uncertain |
| 4021 | NLGN3 | Y | ? | chrX:71167650:71167650:G:A:HEM: NM_181303.2(NLGN3):c.1553G>A p.(Trp518*) | 518 | LOF | N | Inheritance unknown, likely to be DECIPHER 279406 | Likely Pathogenic |
| 4248 | PHIP | Y | ? | chr6:78946244:78946243:-:T:HET:frameshift_variant: Alamut Chr6(GRCh38):g.78946244dup NM_017934.7(PHIP):c.4387dup p.(Arg1463Lysfs*35) g.dupT c.dupA | 1463 | LOF | N | Inheritance unknown, see PMID: 31337854 | Likely Pathogenic |
| 3036 | SETD5 | Y | ? | Chr3(GRCh38):g.9441638del NM_001080517.1(SETD5):c.856del p.(Leu286*) | 286 | LOF | N | Likely clinical diagnosis | Pathogenic |
| 3037 | EP300 | Y | ? | Chr22(GRCh38):g.41177730_41177731del NM_001429.3(EP300):c.6019_6020del p.(Gln2007Valfs*65) | 2007 | LOF | N | Likely clinical diagnosis | Pathogenic |
| 4497 | ARID1B | Y | ? | NM_001346813.1(ARID1B):c.65_66del p.(Glu22Glyfs*209) dn low level mosaic | | | N | De novo | Not LP/P |
| 3046 | NR2F1 | Y | ? | chr5:93585427:93585427:G:A:ENST00000327111.8:c.404G>A; ENSP00000325819.3:p.Arg135His; SIFT: Deleterious (0); PolyPhen: Probably damaging (1); CADD32; REVEL0.962; SpliceAI≤ 0.2 | 135 | MIS | N | Likely clinical diagnosis | Likely Pathogenic |
| 4507 | IFIH1 | Y | ? | chr2:162268237:162268237:A:G:HET:missense_variant:SIFT=deleterious(0.0):PolyPhen =possibly damaging(0.554): | | | Y | Inheritance unknown, implausible MAF | Not LP/P |
| 4507 | RAI1 | Y | ? | chr17:17797774:17797774:C:T:HET:missense_variant:SIFT=deleterious(0.0):PolyPhen=p robably damaging(0.928): | | | Y | Inheritance unknown, implausible MAF | Not LP/P |
| 3188 | EP300 | Y | ? | Chr22(GRCh38):g.41162780G>C NM_001429.3(EP300):c.3728+1G>C p.? | 1243 | ESS | N | Likely clinical diagnosis | Pathogenic |
| 4665 | CASK | Y | ? | chrX:41626688:41626688:C:T:HEM:ENST00000378163.7:c.931G>A; ENSP00000367405.1:p.Ala311Thr; SIFT: Deleterious (0.04); PolyPhen: Benign (0.065); CADD23.7; REVEL0.313; SpliceAI≤ 0.2 | | | N | Inheritance unknown | Not LP/P |
| 3236 | KMT2A | Y | ? | Chr11(GRCh38):g.118484286dup NM_001197104.1(KMT2A):c.4190dup p.(Val1398Serfs*9) | 1398 | LOF | N | Likely clinical diagnosis | Pathogenic |
| 3379 | ANKRD11 | Y | ? | Chr16(GRCh38):g.89280521del NM_013275.5(ANKRD11):c.6021delC p.(Phe2008Serfs*79) | 2008 | LOF | N | Likely clinical diagnosis | Pathogenic |
| 3471 | ANKRD11 | Y | ? | Chr16(GRCh38):g.89284635_89284639del NM_013275.5(ANKRD11):c.1903_1907del p.(Lys635Glnfs*26) | 635 | LOF | N | Likely clinical diagnosis | Pathogenic |
| 4252 | ANKRD11 | Y | Y | Chr16(GRCh38):g.89281638_89281639insGC NM_013275.5(ANKRD11):c.4903_4904insGC p.(Leu1635Argfs*52) | 1635 | LOF | N | Likely clinical diagnosis | Pathogenic |
| 4294 | ANKRD11 | Y | Y | Chr16(GRCh38):g.89282611G>A NM_013275.5(ANKRD11):c.3931C>T p.(Arg1311*) | 1311 | LOF | N | Likely clinical diagnosis | Pathogenic |
| 4348 | ANKRD11 | Y | ? | Chr16(GRCh38):g.89284130_89284134del NM_013275.5(ANKRD11):c.2408_2412del p.(Lys803Argfs*5) | 803 | LOF | N | Likely clinical diagnosis | Pathogenic |
| 4753 | ANKRD11 | Y | Y | Chr16(GRCh38):g.89284364_89284367del NM_013275.5(ANKRD11):c.2175_2178del p.(Asn725Lysfs*23) | 725 | LOF | Y | Looks mosaic in gnomAD, ClinVar have it as LP, one allele in individual >70 yrs ? Benign clonal expansion | Pathogenic |
| 3027 | CACNA1A | Y | ? | chr19:13275893:13275893:C:T; ENSP00000353362.5:p.Asp1316Asn; Sift: Deleterious (0); PolyPhen: Possibly damaging (0.655); CADD27.6; REVEL0.94; SpliceAI≤ 0.2 | | | N | near cluster of LP/P missense, no clinical fit | Not LP/P |
| 3177 | NIPBL | Y | ? | Chr5(GRCh38):g.37036491A>C ENST00000282516.13:c.5971+4A>C; CADD21.2; SpliceAI≤ 0.2 | 1990 | ESS | N | SpliceAI not supportive, inheritance unknown, typical CdLS | Possible |
| 4482 | SET | Y | Y | deletion chr9:128678964-128699851 | | DEL | N | | Likely Pathogenic |
