## Supplementary Table 3 for "Whole genome sequencing of ‘mutation-negative’ individuals with Cornelia de Lange Syndrome"

**Supplementary Table 3 - *NIPBL* non-coding variants identified in individuals affected with CdLS**

| CHROM | POS | REF | ALT | Feature | Variant | Consequence | Kozak | Details | ClinVar ID | ClinVar significance | REF DOI | In this study |
|---|---|---|---|---|---|---|---|---|---|---|---|---|
| 5 | 36876791 | C | T | ENST00000282516 | c.-467C>T | uAUG_gained | Strong | 156bp long uORF | 1195876 | Pathogenic/Likely_pathogenic | 10.3390/genes13050740 | Y |
| 5 | 36876801 | GA | AT | ENST00000282516 | c.-457_-456delinsAT | uAUG_gained | Moderate | 270bp long uORF | 1300231 | Likely_pathogenic | 10.1002/humu.24384 | |
| 5 | 36876937 | CC | A | ENST00000282516 | c.-321_-320delinsA | uORF_frameshift | Moderate | uORF extended from 15 to 189bps | 2151 | Pathogenic | 10.1002/humu.20380 | |
| 5 | 36877164 | C | T | ENST00000282516 | c.-94C>T | uAUG_gained | Weak | 51bp long uORF | NA | | 10.1002/humu.24384; 10.1111/j.1399-0004.2007.00832.x | |
| 5 | 36876936 | GC | G | ENST00000282516 | c.-315del | uORF_frameshift | Moderate | uORF extended from 15 to 189bps | NA | | NA | Y |
