## Supplementary Table 4 for "Whole genome sequencing of ‘mutation-negative’ individuals with Cornelia de Lange Syndrome"

**Supplementary Table 4 - *NIPBL* 5' UTR non-coding variants reported in gnomAD**

| CHROM | POS | REF | ALT | AC | Feature | Variant | Consequence | Kozak | Details |
|---|---|---|---|---|---|---|---|---|---|
| 5 | 36876946 | T | A | 427 | ENST00000282516 | c.-312T>A | uSTOP_lost | Moderate | |
| 5 | 36876976 | G | A | 1 | ENST00000282516 | c.-282G>A | uAUG_gained | Weak | 117bp long uORF |
| 5 | 36876984 | A | C | 1 | ENST00000282516 | c.-274A>C | uAUG_lost | Weak | |
| 5 | 36876984 | A | G | 1 | ENST00000282516 | c.-274A>G | uAUG_lost | Weak | |
| 5 | 36876994 | AGGAG | A | 2 | ENST00000282516 | c.-262_-259del | uORF_frameshift | Weak | uORF extended from 90 to 108bps |
| 5 | 36877040 | G | A | 1 | ENST00000282516 | c.-218G>A | uSTOP_gained | Weak | |
| 5 | 36877059 | T | A | 1 | ENST00000282516 | c.-199T>A | uAUG_lost | Weak | |
| 5 | 36877073 | A | AG | 1 | ENST00000282516 | c.-183dup | uORF_frameshift | Weak | uORF reduced from 66 to 39bps |
| 5 | 36877097 | CA | C | 1 | ENST00000282516 | c.-160del | uORF_frameshift | Weak | uORF extended from 66 to 141bps |
| 5 | 36877098 | A | AC | 7 | ENST00000282516 | c.-153dup | uORF_frameshift | Weak | uORF extended from 66 to 165bps |
| 5 | 36877098 | AC | A | 7 | ENST00000282516 | c.-153del | uORF_frameshift | Weak | uORF extended from 66 to 141bps |
| 5 | 36877118 | T | TTATA | 3 | ENST00000282516 | c.-139_-136dup | uORF_frameshift | Weak | uORF extended from 66 to 168bps |
| 5 | 36877123 | G | C | 1 | ENST00000282516 | c.-135G>C | uSTOP_lost | Weak | |
| 5 | 36877172 | C | CAT | 1 | ENST00000282516 | c.-86_-85insAT | uAUG_gained | Weak | 48bp long uORF |
| 5 | 36953628 | A | G | 2 | ENST00000282516 | c.-69A>G | uAUG_lost | Strong | |
