## Supplementary Figures for "Whole genome sequencing of ‘mutation-negative’ individuals with Cornelia de Lange Syndrome"

Figure S1. IGV plots of *NIPBL* coding regions variants of probands and parents (where available)

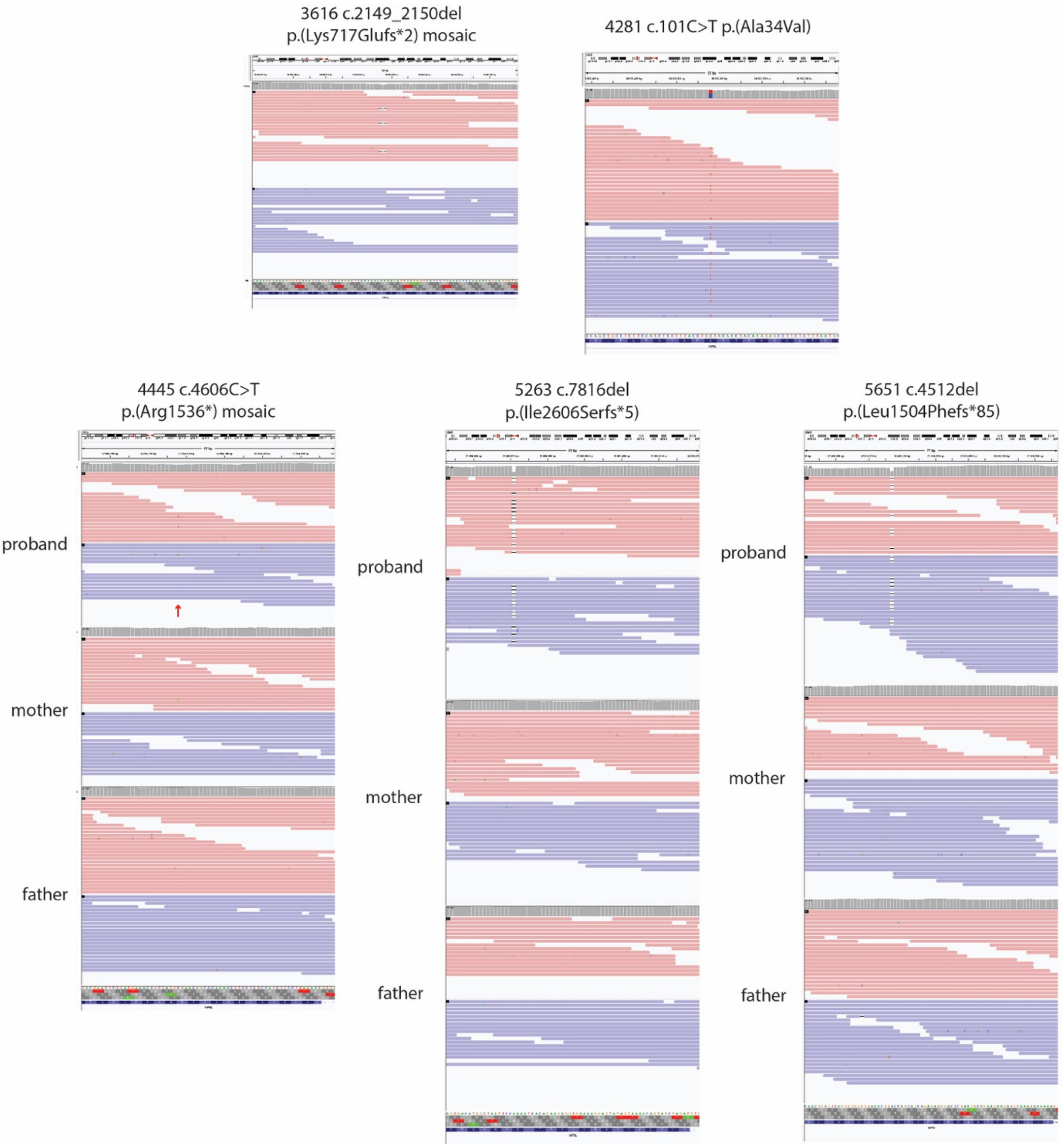

Figure S2. IGV plots of *NIPBL* essential splice site  variants of probands and parents (where available)

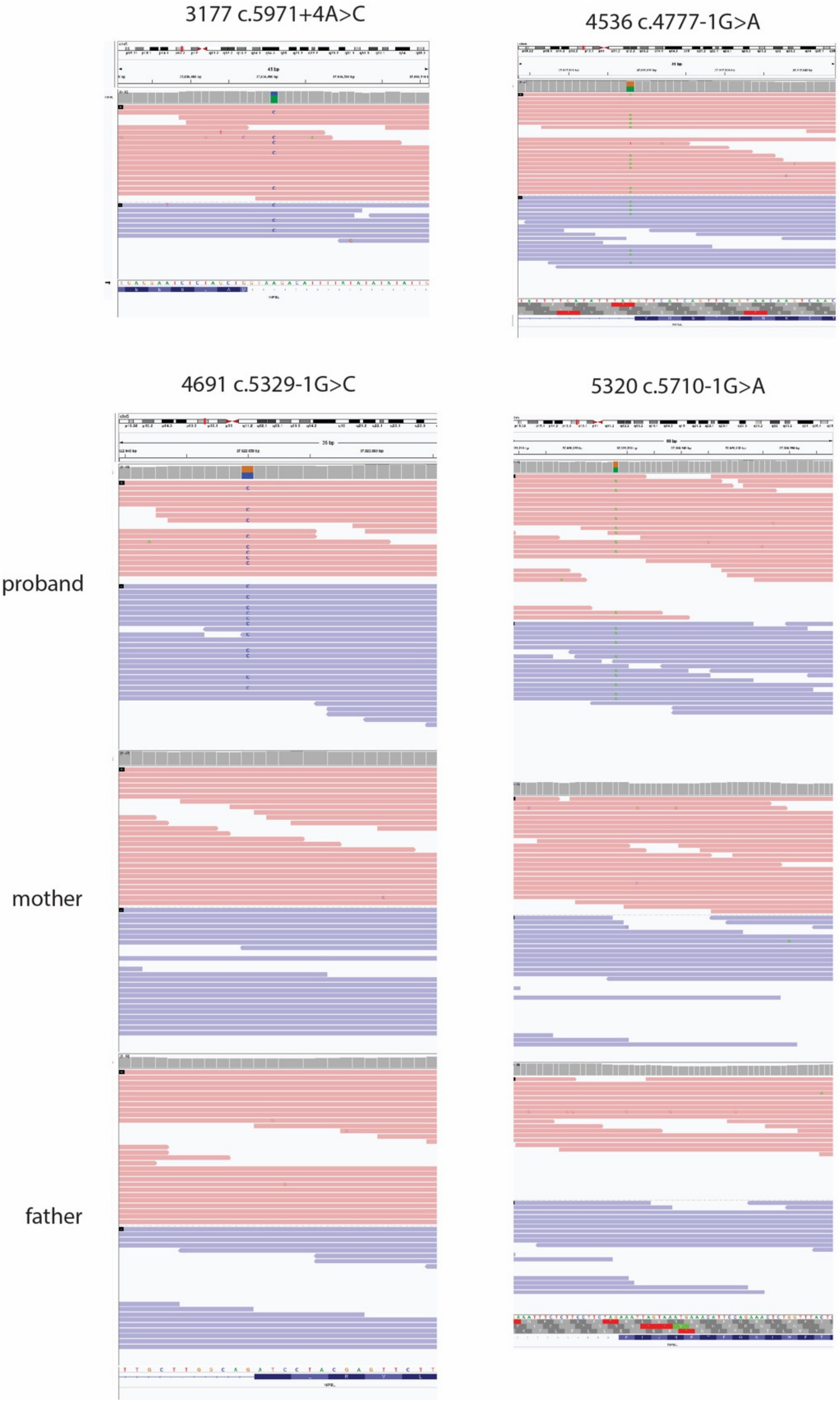

Figure S3. IGV plots of *ANKRD11* coding regions variants of probands and parents (where available)

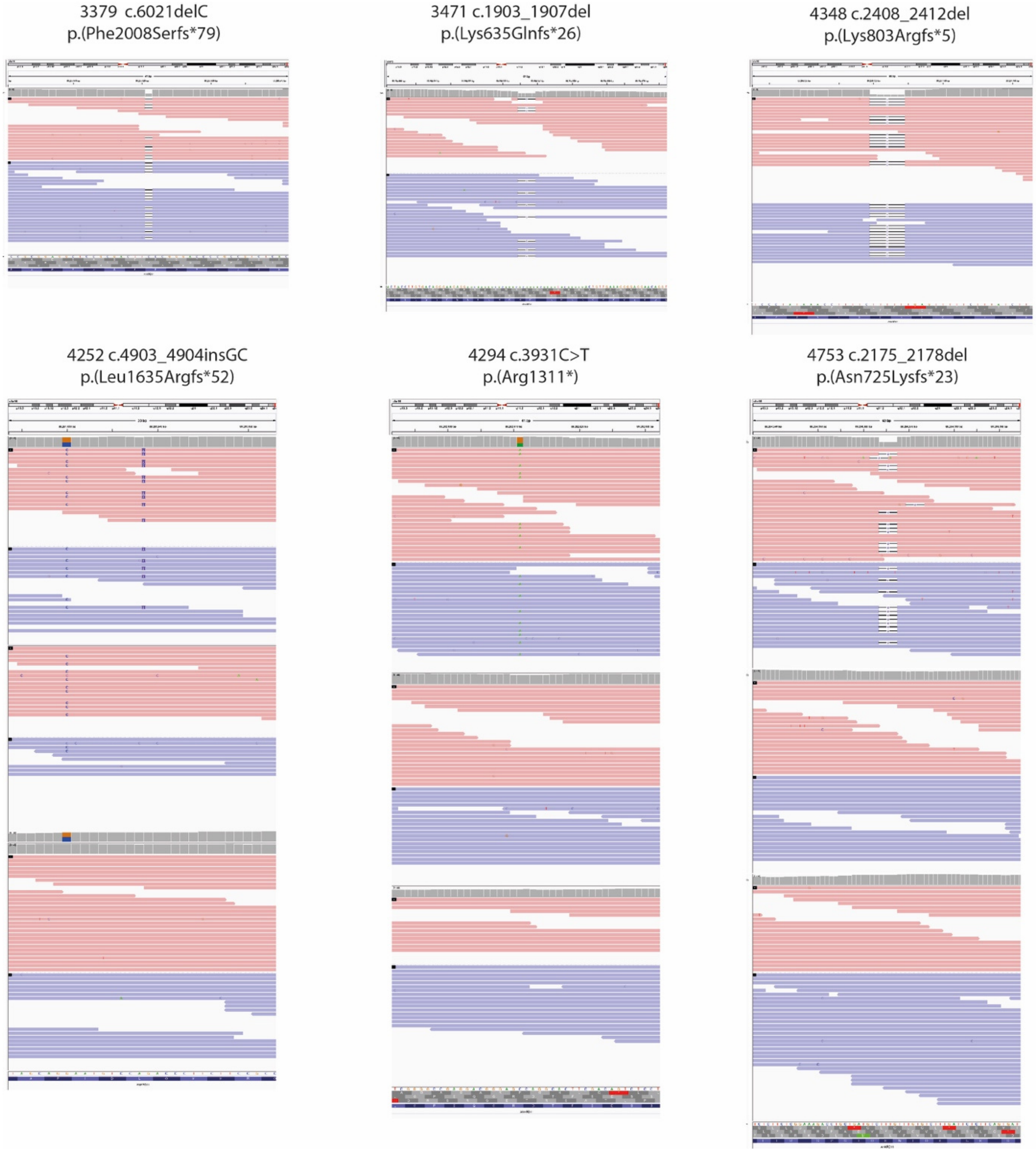
